# Patterns of Failure for Recurrent Head and Neck Squamous Cell Carcinoma Treated with Salvage Surgery and Postoperative IMRT Reirradiation

**DOI:** 10.1101/2023.05.13.23289864

**Authors:** Abdallah S. R. Mohamed, Geoffrey V. Martin, Sweet Ping Ng, Vinita Takiar, Beth M. Beadle, Mark Zafereo, Adam S. Garden, Steven J. Frank, C. David Fuller, G. Brandon Gunn, William H. Morrison, David I. Rosenthal, Jay Reddy, Amy Moreno, Anna Lee, Jack Phan

## Abstract

**Purpose/Objectives:** Determining the optimal postoperative reirradiation volume following salvage surgery for recurrent head and neck squamous cell cancer (HNSCC) is critical given the significant risk of severe reirradiation toxicity and need to provide durable local control. The purpose of this study was to evaluate patterns of locoregional recurrence (LRR) after surgical salvage and adjuvant reirradiation with IMRT.

**Materials/Methods:** Patterns of LRR for 61 patients treated consecutively between 2003-2014 who received post-operative IMRT reirradiation to ≥60 Gy for recurrent HNSCC were determined by 2 methods: 1) physician classification via visual comparison of post-radiotherapy imaging to reirradiation plans; and 2) using deformable image registration (DIR). Those without evaluable CT planning image data were excluded. All recurrences were verified by biopsy or radiological progression. Failures were defined as in-field, marginal, or out-of-field. Logistic regression analyses were performed to identify predictors for LRR.

**Results:** A total of 55 patients were eligible for analysis and 23 (42%) had documented LRR after reirradiation. Location of recurrent disease prior to salvage surgery (lymphatic vs. mucosal) was the only significant predictor of LRR after post-operative reirradiation (p = 0.037). Physician classification of LRR yielded 14 (61%) in-field failures, 3 (13%) marginal failures, and 6 (26%) out-of-field failures, while DIR yielded 10 (44%) in-field failures, 4 (17%) marginal failures, and 9 (39%) out-of-field failures. Most failures (57%) occurred within the original site of recurrence or first echelon lymphatic drainage. Of patients who had a free flap placed during salvage surgery, 56% of failures occurred within 1 cm of the surgical flap.

**Conclusion:** The majority of locoregional recurrences after surgical salvage and adjuvant reirradiation for recurrent HNSCC occurred marginally or in-field, often in or near the anastomotic site.

## INTRODUCTION

Locoregional recurrence (LRR) remains a significant challenge in the management of patients with head and neck squamous cell carcinoma (HNSCC) who have undergone curative radiation therapy (RT). (1) Surgical salvage is a viable option for eligible patients, and adjuvant reirradiation (re-RT) has been shown to enhance locoregional control, especially in those with pathological risk factors such as positive margins, lymphovascular space invasion, perineural invasion, and/or extracapsular extension. However, patients with one or more of these risk factors still face a 40-80% risk of LRR within two years after salvage surgery alone. (2, 3) Consequently, adjuvant re-RT is recommended despite the potential for severe acute and long-term toxicity, with rates of grade 3 or higher (G3+) toxicity reaching up to 60%.(4-6)

To minimize re-RT toxicity risk, treatment target volumes are typically confined to the tumor bed, with little or no prophylactic extension to subclinical disease volumes and/or elective nodal irradiation.(6, 7) Advanced radiotherapy techniques, such as intensity modulated radiation therapy (IMRT) and proton beam therapy (PBT), have emerged as the standard for enhancing normal tissue sparing. Our published studies of head and neck reirradiation using IMRT (n=227) and PBT (n=60) demonstrated 2- and 5-year G3+ rates of 26-32% and 40-48%, respectively.(6, 7) Multivariate analyses revealed that the most significant factor impacting G3+ toxicity was retreatment volume (>50 cc), which supports the use of smaller field sizes to mitigate treatment complications. Nevertheless, 2- and 5-year locoregional control rates among HNSCC patients treated with surgery and adjuvant reirradiation remain suboptimal, at 60% and 50%, respectively. (6, 7)These figures are consistent with several published studies that report inferior outcomes following reirradiation among HNSCC patients.(8, 9) This underscores the need for optimization of reirradiation tumor volumes. For example, an in-field local recurrence implies a problem related to tumor radioresistance and dose-optimization, while a marginal recurrence suggests a geometric miss and the necessity for adequate target coverage.

Despite the clear need for locoregional pattern of failure (POF) analyses after reirradiation for HNSCC, such studies have not been rigorously conducted or assessed using deformable image registration (DIR) techniques.(10-12) Our group developed a unique methodology to standardize the analysis and reporting of HNSCC patterns of failure using both geometric and dosimetric parameters.(13) We previously implemented this methodology for POF analysis in different HNSCC sites after primary RT.(14-17) In the current work, we aim to evaluate this methodology in recurrent/second primary HNSCC patients who have undergone salvage surgical resection and IMRT reirradiation. Our goal is to optimize target delineation and treatment volume for HNSCC reirradiation, ultimately improving patient outcomes.

## METHODS

### Patient selection

The charts of patients who received IMRT reirradiation to the head and neck from 2003-2014 at our institution were retrospectively reviewed under an IRB approved protocol (PA12-0168). Patients were eligible for this study if they had prior history of radiation to the head and neck >30 Gy, underwent salvage surgery as initial treatment for their recurrent disease, and had pathologically confirmed HNSCC arising from the oral cavity, oropharynx, nasopharynx, salivary glands, paranasal sinuses, or larynx/hypopharynx. If patients had multiple courses of salvage surgery followed by reirradiation only the first reirradiation course was considered in the analyses. Only patients who received a prescribed reirradiation dose of at least 60 Gy and began reirradiation therapy within 90 days of salvage surgery were included.

### Reirradiation Plans

Although there were no standard definitions used for re-RT volume delineation, the clinical target volume (CTV1) typically included the entire tumor bed with a margin (0.8-1.0 cm) and a subsequent CTV to cover the operative bed, at risk subclinical regions and/or reduced-volume elective nodal region (CTV2, CTV3) at the discretion of the treating radiation oncologist. All contours and patient records were reviewed biweekly at our Quality Assurance Head and Neck Radiation Oncology Planning Conference, which included physical examination, as previously described.(18, 19)

The re-RT plans for the patient’s included in this study were analyzed in the Pinnacle treatment planning software. All radiation plans had at least 2 prescribed dose levels with at least one of those dose levels prescribed to ≥60 Gy (CTV1). The second (or third) dose levels (CTV2, CTV3) were prescribed to doses of <60 Gy but ≥50 Gy for coverage of elective neck nodal disease. All reirradiation plans were administered with the use of IMRT with the aforementioned target volumes and additional organs at risk delineated by the treating physician.

### Pattern of Failure Analyses

Figure 1 depicts a flow diagram used for patients selected for the final analyses. All failures after re-RT had post-treatment CT/PET-CT imaging depicting the location of failure and were either pathologically confirmed or showed evidence of radiographic progression on ≥2 post-treatment imaging studies. The date of failure was designated at the first radiographic finding of recurrence or biopsy confirmation.

**Figure 1.**
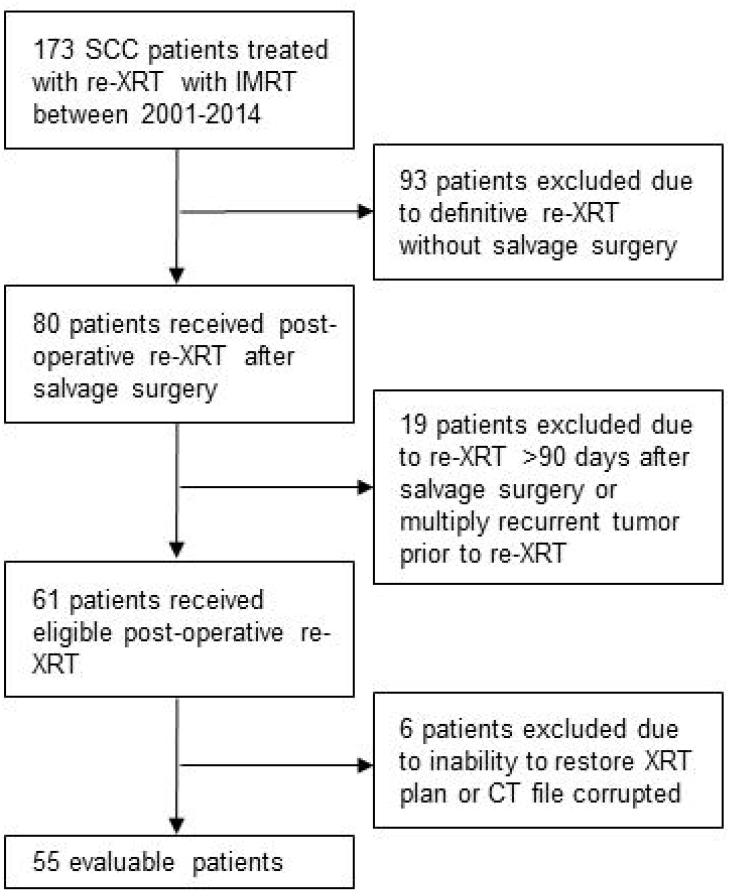
Flow diagram depicting patients selected for inclusion in study. XRT: radiation therapy, IMRT: intensity modulated radiation therapy, CT: computed tomography

LRR was defined as any recurrence in non-central nervous system sites of the head or neck superior to the clavicles, including the contralateral head/neck, lymphatics, mucosa of the aerodigestive tract, paranasal sinuses, soft/subcutaneous tissue, and bone. Distant failures were defined as any which did not meet these criteria.

The type of LRR was defined based on their location relative to the reirradiation field and defined as in-field, marginal, and out-of-field (see below). The clinical and pathologic features were analyzed to identify predictors of LRR.

We used two different methods to determine POF. The first method involved visual comparison of axial slices between the post re-RT diagnostic scans depicting recurrence (diagnostic rCT) and the IMRT reirradiation therapy plan containing isodose curves. A local recurrence was defined as in-field if the centroid of recurrence (i.e., center of the 3-dimensional tumor recurrence volume) was within CTV1 and marginal if not in-field but within CTV2-3. An out-of-field recurrence was defined as not meeting the definition of a local recurrence (centroid of recurrence outside CTV1-3). The type of LRR was independently classified by three head and neck physicians (2 radiation oncologists, 1 head and neck surgeon). The results were pooled and the interobserver variability was assessed.

The second method of POF analysis utilized DIR of image datasets acquired from the diagnostic rCT and IMRT reirradiation planning CT. Specifically, the tumor recurrence on the diagnostic rCT was delineated by a radiation oncologist. This structure was then propagated onto the IMRT reirradiation planning CT. Deformable registrations were performed using Velocity software (Varian Medical Systems, Palo Alto, CA). The type of LRR was then defined as either in-field, marginal, or out-of-field based on centroid of recurrence using the same definitions listed above. Further stratification of DIR LRR type was performed by spatial assessment of the centroid of recurrence relative to the prescription CTV isodose lines based on the dose to 95 percent of the recurrence volume (D95). This sub-classification yielded 5 types of recurrences (Type A-E). The in-field and marginal recurrences were each split into 2 subcategories based on D95 dose: in-field Type A (D95≥60 Gy) and C (D95<60 Gy); and marginal Type B (D95≥54 Gy) and D, (D95<54 Gy)). Type E was defined as recurrence completely outside the prescription isodose lines for all CTVs (CTV1-3). This methodology has been described in prior publications from our institution.(13)

### Statistical Analyses

Ordinal variables are summarized by the number (%) of occurrences and continuous variables are summarized as the median (range) unless otherwise specified in the text. Baseline and salvage patient and treatment characteristics, including age, gender, location of primary tumor which received 1^st^ course of radiation therapy, site of recurrence which underwent salvage surgery and reirradiation, reirradiation volume, pathologic, and surgical information were evaluated as covariates of LRR after reirradiation. Univariate logistic regression was performed and statistically significant variables were included in a multivariate model. POF analysis was summarized by visual and DIR methods with interobserver variability determined by interclass correlation. Statistical analyses were performed with SPSS version 23 (Armonk, NY). *P* < 0.05 were considered statistically significant.

## RESULTS

### Patient Characteristics

Patient demographic details and salvage treatment characteristics are listed in Table 1. In total 61 patients met the criteria for adjuvant reirradiation and of these 55 (90%) had evaluable CT planning data for final analysis (Figure 1). Median follow-up of patients was 22 months (range 1-105 months). The median time between the original radiation course and first recurrence was 21 months (range 3-385 months). The site of recurrence prior to salvage surgery was further aggregated as a mucosal (oral cavity, oropharynx, nasopharynx, sinonasal, hypopharynx, and larynx) vs. neck (lymphatic/skin), with 33% of patients having a mucosal only recurrence, 60% having a neck only recurrence, and 7% having a recurrence in both the mucosa and neck.

**Table 1.** Patient Characteristics

| Characteristic | LR<br>Recurrence | No LR<br>Recurrence |
| --- | --- | --- |
| N | 23 | 32 |
| Age at reirradiation (years), median (range) | 64 (27-84) | 60 (35-82) |
| Gender (% male) | 74% | 81% |
| Location of original tumor, n (%) |  |  |
| Nasopharynx | 0(0%) | 2 (6%) |
| Skin/scalp | 2 (9%) | 2 (6%) |
| Oropharynx | 4 (17%) | 4 (13%) |
| Oral cavity | 7 (30%) | 8 (25%) |
| Larynx/Hypopharynx | 7 (30%) | 10 (31%) |
| Sinonasal | 2 (9%) | 3 (9%) |
| Salivary/Other | 1 (4%) | 3 (9%) |
| Recurrence location prior to salvage surgery |  |  |
| Mucosal only, n (%) | 12 (52%) | 6 (19%) |
| LN/soft tissue/skin only, n (%) | 11 (44%) | 22 (68%) |
| Both mucosal and LN, n (%) | 1 (4%) | 4 (13%) |
| Time to first recurrence after original radiation (months),<br>median (range) | 15 (3-235) | 22 (3-385) |
| Surgery at time of primary radiation, n (%) | 6 (26%) | 7 (22%) |
| Salvage surgery free flap use, n (%) | 16 (70%) | 15 (47%) |
| Pathological Features |  |  |
| Mucosal recurrences |  |  |
| Size of tumor (cm), median (range) | 1.9 (0.8-6.1) | 3.0 (0.9-5.5) |
| PNI, % positive | 87% | 60% |
| Margins, % positive (%close) | 47% (7%) | 10% (20%) |
| LN/soft tissue/skin recurrence |  |  |
| ECE, % positive | 83% | 96% |
| Number of LN positive, median (range) | 1 (1-9) | 1 (1-11) |
| Total radiation dose (Gy), median (range) | 126 (100-138) | 128 (114-142) |
| Reirradiation CTV1 mean dose (Gy), median (range) | 62 (61-72) | 62 (34-92) |
| Reirradiation CTV1 volume (mL), median (range) | 78 (7-735) | 72 (9-401) |
| Chemotherapy with reirradiation (% yes) | 70% | 84% |
| Induction, n (% total patients) | 3 (13%) | 1 (3%) |
| Concurrent, n (% total patients) | 12 (43%) | 17 (53%) |
| Induction and Concurrent, n (% total patients) | 3 (13%) | 8 (25%) |
| Adjuvant and concurrent, n (% total patients) | 0 (0%) | 1 (3%) |

LR: locoregional, LN: lymph node, PNI: perineural invasion, ECE: extracapsular extension

Surgical reconstruction was performed with a free flap in 56% of patients, with the remaining patients having anastomosis/primary closure. The median CTV1 reirradiation dose was 60 Gy for all patients, and the mean CTV1 reirradiation volume was 75 cc. Six patients (11%) received prescription reirradiation doses >60 Gy. Chemotherapy in the induction, concurrent, or adjuvant setting was used in conjunction with reirradiation in 82% of patients, with the majority of patients receiving concurrent chemotherapy.

### POF Analysis

Of the 55 evaluable patients, 23 had LRR with a median time to recurrence of 5 months (range 1-16). Fifteen patients had a distant failure, with 7 patients presenting with both LRR and distant failure. Among the 23 LRR cases, 9 (40%) recurred in the same subsite as the initial recurrence and 4 (17%) recurred in the 1^st^ echelon lymph node level of the initial recurrence. Of the remaining 10 (43%) recurrences, 6 (60%) were mucosal recurrences >2 cm from the reirradiation volume, and 4 (40%) were nodal recurrences not in the normal drainage pattern of the primary recurrence. LRR details of each case are listed in Table 2.

**Table 2.**
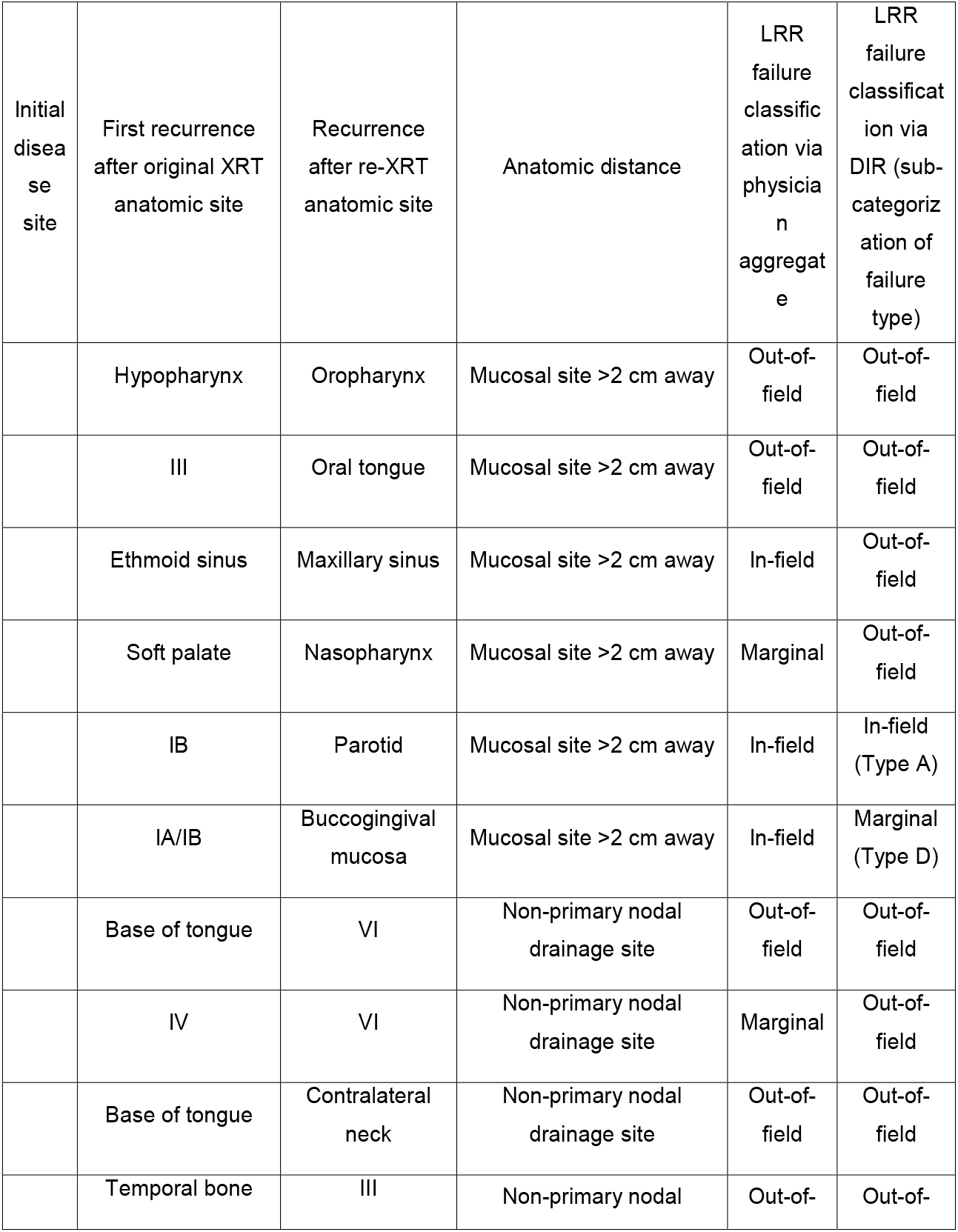
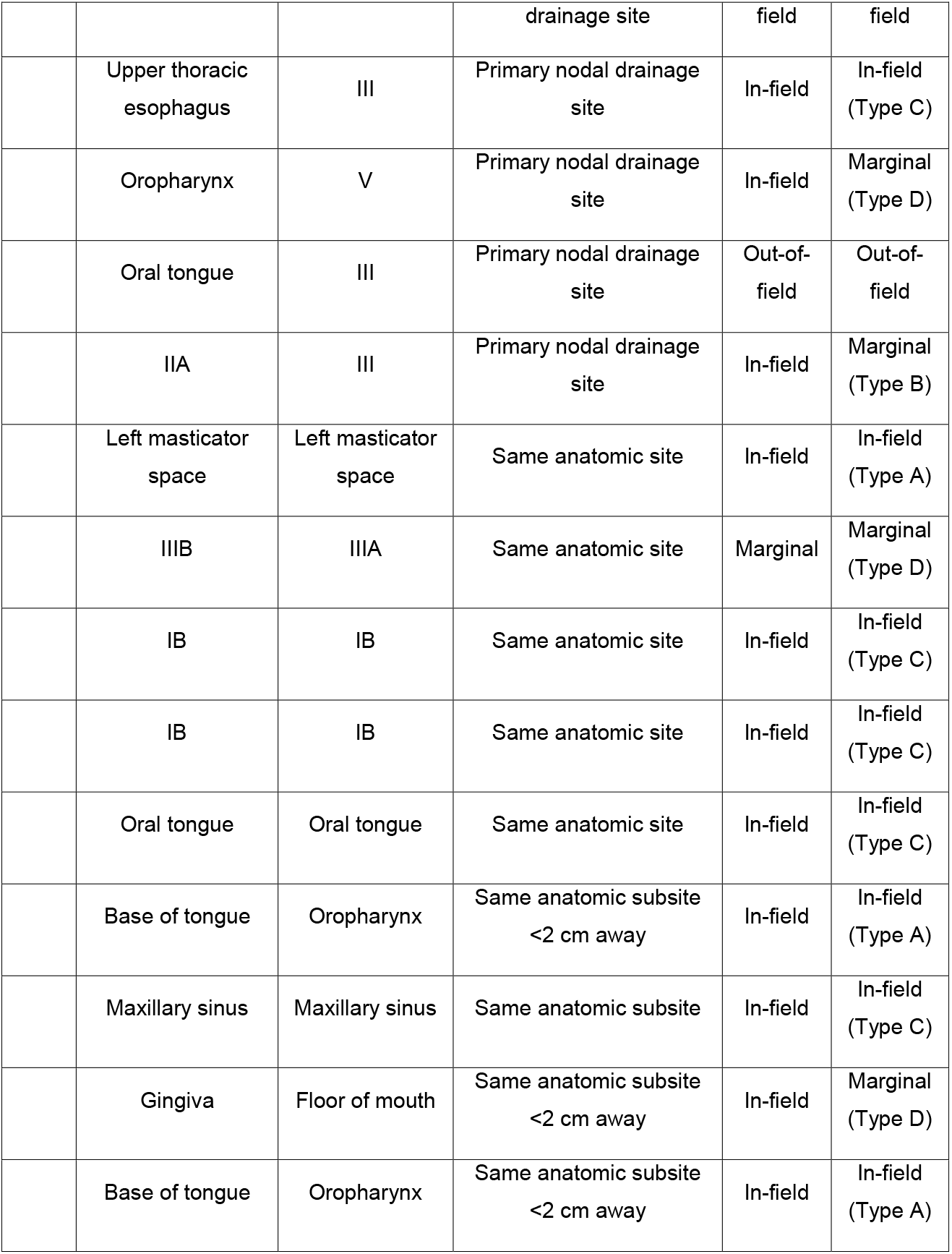
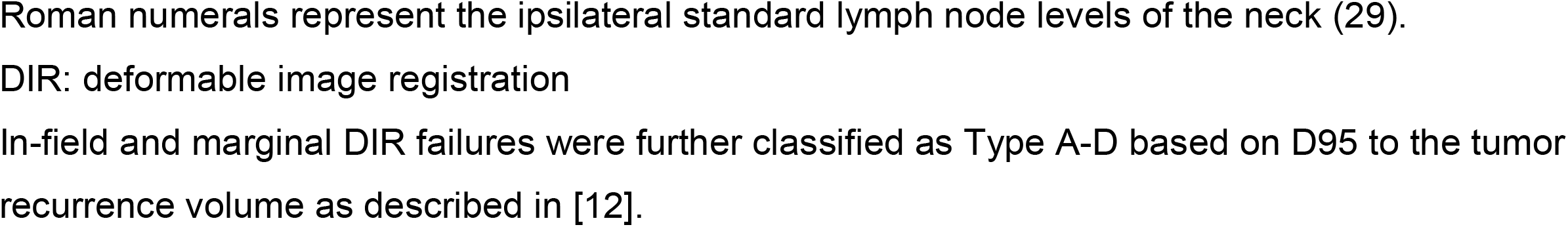
Anatomic distance of locoregional failures after post-operative reirradiation

Baseline patient and salvage treatment factors were analyzed as univariate predictors of LRR. Significant predictors were site of recurrence (mucosal vs neck; p = 0.037) and the presence of PNI (p = 0.047). Margin status, extracapsular extension, presence of LVSI, patient age, gender, CTV1 max dose, CTV1 volume, and original treatment site were not predictive of LRR, consistent with our prior report.(6)

POF analysis by visual inspection (i.e., comparison of radiation treatment plans and post-radiation surveillance imaging) for each physician observer is displayed in Figure 2. All cases (23 of 23) had at least 2 physicians agree on the POF classification, 52% (12 of 23) of cases had unanimous agreement among all 3 physicians. Final POF classification (as determined by agreement between at least 2 physicians) demonstrated that 74% (17 of 23) of recurrences were local and 26% (6 of 23) were out-of-field. Of the 17 local recurrences, 15 (88%) were classified as in-field and 2 (12%) as marginal. Interobserver agreement demonstrated an interclass correlation of 0.68.

**Figure 2.**
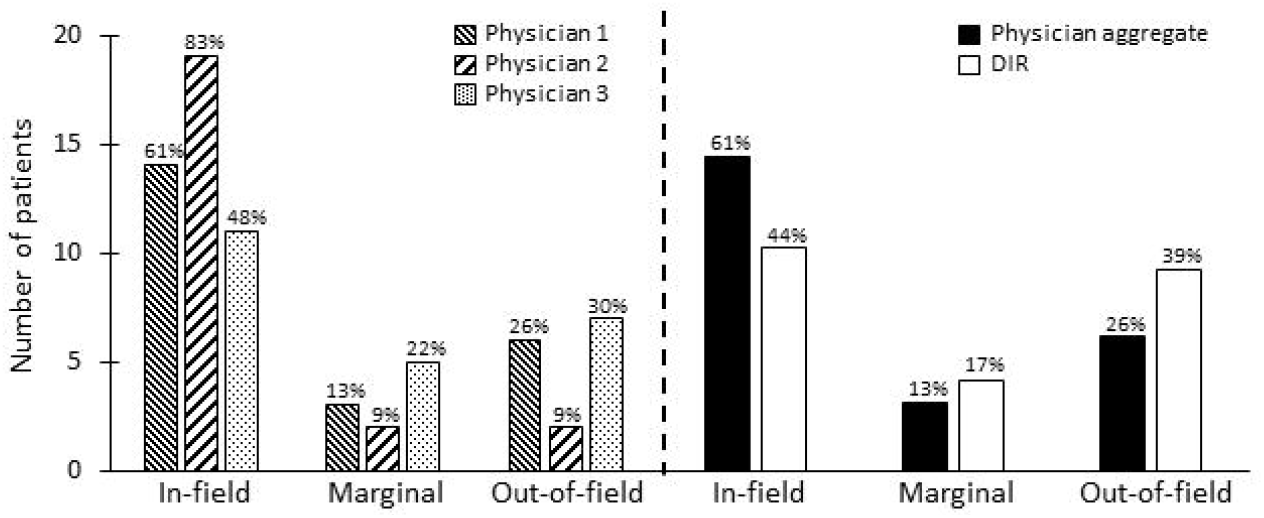
Classificationof locoregional failure types by physician assignment and deformable image registration (DIR) methods. Locoregional failures were classified as in-field, marginal, or out-of-field using these 2 methods. The left panel depicts failure classifications by each individual physician, and the right panel depicts the aggregate physician failure classification type vs. assignment by deformable image registration. The percentage of patients who experienced each locoregional failure type as determined by the different methods is noted above each column.

POF analysis by DIR classified 61% (14 of 23) as local failures and 39% (9 of 23) as out of field failures. Among local failures 64% (9 of 14) were in-field and 36% (5 of 14) were marginal. Further stratification of the 9 in-field recurrences revealed that 4 (44%) were within the high dose (D95 ≥ 60 Gy) region and 5 (56%) were within the intermediate dose (D95 < 60 Gy) region.

Comparison between visual inspection and DIR POF analyses showed 6 cases reclassified by DIR. Two marginal failures by visual inspection were reclassified as out of field by DIR; 3 in-fields were reclassified as marginal by DIR; and 1 in-field reclassified as out-of-field by DIR (Table 2). An example of 2 reclassified patients is depicted in Figure 3.

**Figure 3.**
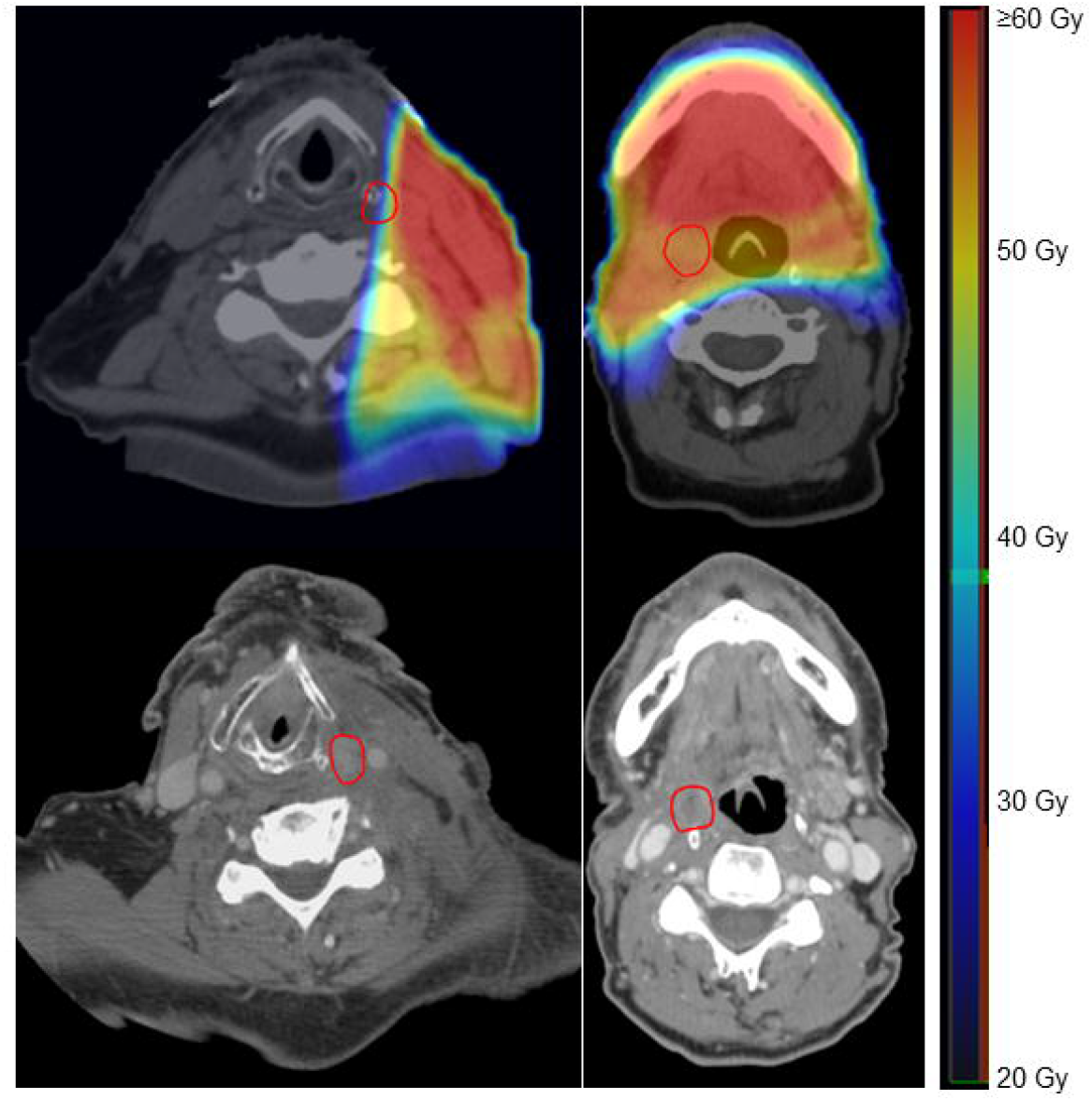
Examples of DIR registrations between re-irradiation simulation CT and recurrence CT scans. The top panels represent 2 patients’ re-irradiation simulation CT and plan with recurrence contour (red circle) propagated from recurrence CT (bottom panels) using deformable image registration. Axial sliceswere taken at the centroid of tumor recurrence. Patient on the left was classified as out-of-field recurrence via DIR and marginal recurrence by physicians via visual inspection, while the patient on the right was classified as marginal via DIR and in-field by physicians. CT: computed tomography.

Analysis by anatomic location demonstrated that after reirradiation, 40% of tumors recurred in the same subsite as the initial recurrence, 17% recurred in the 1^st^ echelon lymph node level of the initial recurrence, and 43% recurred outside of these anatomic zones (Table 2). Additionally, 56% of tumors (9 of 16) recurred within 1 cm of the flap reconstruction bed (Figure 4). Of these, 2 were in-field and 7 were marginal by DIR method. In terms of coverage, 16 of 23 (70%) had the entire surgical bed was covered by ≥ 54 Gy, while 7 did not receive full coverage of the surgical bed by any prescription dose. Among the 7 recurrences without full tumor bed coverage, 1 was in-field, 1 was marginal, and 5 were out-of-field.

**Figure 4.**
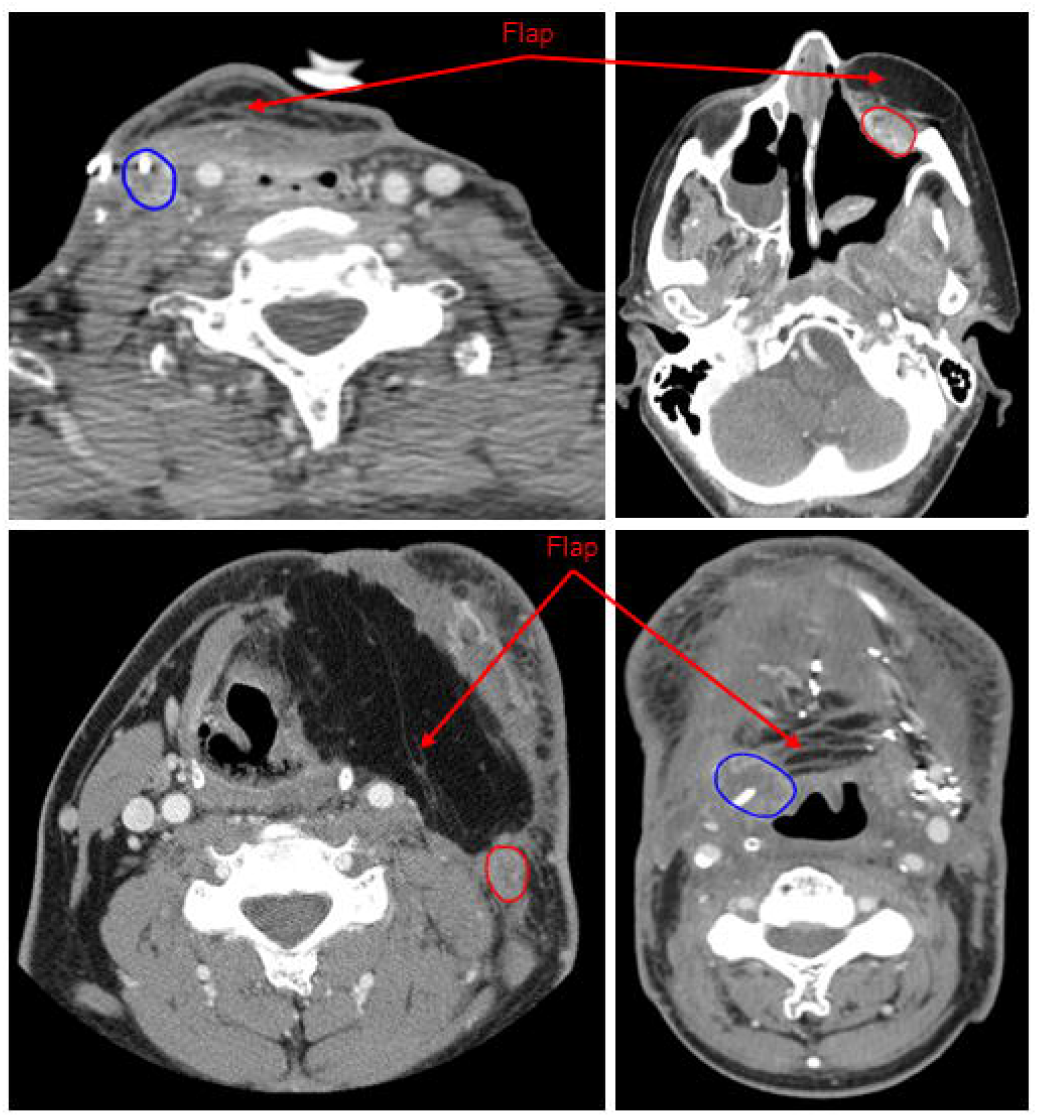
Examples of 4 locoregional failures after re-irradiation near surgical flaps. Images represent the contrast enhanced CT which detected a head and neck tumor recurrence. Recurrent tumors are circled in blue/red and red arrows point to the location of the surgical flap on the recurrence CT scans. CT: computed tomography.

## DISCUSSION

The results from this study demonstrate that the majority of LRR for recurrent HNSCC after salvage surgery and IMRT reirradiation were within or near the reirradiation treatment field. These results were generally consistent between visual inspection and DIR techniques, with the exception of more local failures classified as marginal rather than in-field when using DIR compared to visual. A major predictor of LRR following salvage surgery and adjuvant reirradiation was the site of first recurrence (mucosal vs neck), with two-thirds of neck only recurrences salvageable with surgery and postoperative reirradiation. Furthermore, most local recurrences occurred within the primary reirradiation site or within the primary lymphatic drainage echelon and were commonly adjacent to the surgical flap.

The pattern of failure outcomes reported in this study are commensurate with other studies evaluating LRR POF after salvage surgery and reirradiation. For example, a report of 42 patients from the Mayo Clinic demonstrated a 54% rate of LRR within the reirradiation field, while a study of 39 patients by Kasperts et al observed that all LRR occurred in the high dose volume (10, 20). In contrast, Margalit et al. demonstrated that >70% of LRR were out-of-field in the post-operative setting, with marginal failures occurring only in patients who did not receive complete reirradiation coverage of the post-operative bed (21). Studies which have included both post-operative and definitive reirradiation in their POF analyses also support high rates of in-field recurrences of >80%.(11, 12) Given these findings, there has been interest in treatment intensification to improve local control for recurrent HNSCC. Early reirradiation studies have associated an improvement in local outcomes with reirradiation doses of >50 Gy.(22) A phase I trial of bevacizumab, 5-FU, and hydroxyurea along with a median radiation dose of ≥66 Gy for high risk head and neck cancer patients demonstrated a <25% in-field recurrence rate in the reirradiation setting.(23) Our group also recently reported that reirradiation doses of ≥70 Gy and concurrent chemotherapy in those with unresectable disease receiving definitive reirradiation were associated with better locoregional control. In the current study cohort, the DIR technique demonstrated that ∼50% of in-field recurrences had a D95 that was below the prescribed high dose (60 Gy).

A major goal of this study was to determine the optimal reirradiation volume. In contrast to the primary treatment of HNSCC where radiation volumes are expanded to include at-risk lymphatics and areas of microscopic disease risk, the reirradiation volumes utilized at our institution are typically limited to the highest risk areas as a tradeoff between cancer control and significant toxicity. This approach is consistent with the majority of postoperative reirradiation studies, including Janot et al that generally prescribed the high dose volume to a 1-2 cm expansion around the gross tumor volume or tumor bed, and sparingly used prophylactic nodal irradiation volumes.(1, 10, 20, 24) Utilizing DIR, we demonstrated the majority of failures (57%) occurred within 2 cm of the initial recurrence subsite or first echelon neck nodes. Among those who had a free flap reconstruction and flap recurrence, 60% recurred within 1 cm of the flap. In addition, a higher percentage of out-of-field failures were observed when the entire surgical bed was not covered in the radiation field. These findings are consistent with prior studies demonstrating a high percentage of local failures occur at the edge of the surgical flap.(25, 26) This may be attributed to the pro-inflammatory, growth factor rich environment in these regions (27). Particular attention should be paid to the primary site of recurrence with attention to the salvage surgical bed and site of flap anastomoses, along with consideration of the primary lymphatic echelon when determining high risk regions during treatment planning.

Our group previously published on the benefits of using DIR techniques compared to rigid registration with regards to CT imaging registration quality in the head and neck.(28) The taxonomy for classifying head and neck failures after IMRT developed from these studies formed the basis of failure definitions used in the current study.(13) This approach takes into account both dose to the centroid of recurrence and to 95% of the recurrence volume.

Compared to physician assigned failure definitions based on visual comparison of IMRT isodose lines and recurrence imaging, DIR resulted in more failures defined as marginal/out-of-field. However, in the majority of cases, this difference, as demonstrated by Figure 2, may not represent distinct biologic subtypes given that the D95 of almost all recurrences was <60 Gy. To our knowledge this is the first study to compare physician assigned and DIR failure classification methods after reirradiation, and additional research is needed to further refine differences in marginal/out of field treatment failures.

As with any retrospective study, the current investigation suffers from a few limitations. Specifically, the patients included in this study represent a heterogeneous group with the possibility of selection bias. While we did our best to analyze a homogenous population by including patients who received only IMRT, SCC histology, had gross total resection, and reirradiation within 90 days of surgery, the potentially biologically distinction of mucosal vs. nodal recurrences were not assessed. Similarly, factors related to systemic therapy and chemotherapy timing, time to initial recurrence after primary radiotherapy and/or reirradiation may also impact POF behavior. In addition, some patients included in this analysis received their original course of radiation >15 years ago and did not have available DICOM files from the primary radiation, precluding registration of these radiation fields to the reirradiation plans for a more comprehensive evaluation of POF. Nevertheless, despite these limitations, this is one of the largest studies investigating the patterns of failure in a cohort of patients who received post-operative reirradiation for recurrent head and neck cancer.

In conclusion, we have shown the majority of LRR after salvage surgery and reirradiation for HNSCC occur in or near the prescribed reirradiation field, and typically within the same anatomic subsite or first echelon nodal station. In addition, the most significant factor predicting LRR after reirradiation is the site of the recurrence prior to salvage surgery and other treatment aspects appear less important. A prospective study to identify patients who would benefit most from the combination of salvage surgery and reirradiation and the optimal reirradiation volume to improve the therapeutic ratio in this very high-risk population is warranted.

## Data Availability

All data produced in the present study are available upon reasonable request to the authors

